# Implementing *Essential Coaching for Every* Mother during COVID-19: A Pilot Pre-Post Intervention Study

**DOI:** 10.1101/2021.01.13.21249598

**Authors:** Justine Dol, Megan Aston, Amy Grant, Douglas McMillan, Gail Tomblin Murphy, Marsha Campbell-Yeo

## Abstract

**Objectives:** The primary objective was to evaluate the preliminary impact of *Essential Coaching for Every Mother* on self-efficacy, social support, postpartum anxiety and postpartum depression. The second objective was to explore the acceptability of the *Essential Coaching for Every Mother* program provided during the COVID-19 pandemic.

**Methods:** A prospective pre-post study was conducted with first time mothers in Nova Scotia, Canada between July 15^th^ and September 19^th^, 2020. Participants completed a self-report survey at enrolment (after birth) and six-weeks postpartum. Various standardized measures were used and qualitative feedback on the program was also collected. Paired t-tests were carried out to determine changes from baseline to follow-up on psychosocial outcomes and qualitative feedback was analysed through thematic analysis.

**Results:** A total of 88 women enrolled. Self-efficacy increased between baseline (B) and follow-up (F) (*B*:33.33; *F*:37.11, *p*=0.000) while anxiety (STAI) declined (*B*:38.49; *F*:34.79; p=0.004). In terms of acceptability, 89% of participants felt that the number of messages were just right, 84.5% felt the messages contained all the information they needed relative to caring for a newborn and 98.8% indicated they would recommend this program to other new mothers.

**Conclusion:** *Essential Coaching for Every Mother* may play a role in increasing maternal self-efficacy and decreasing anxiety, although future work with a control group is needed to delineate the true effects of the program. Overall, mothers were satisfied with the *Essential Coaching for Every Mother* program and would recommend it for other mothers, during COVID-19 and beyond.

## Introduction

In the postpartum period, women face challenges in the ability to access information and find adequate support,^1^ which has only been magnified during the COVID-19 pandemic.^2^ In March 2020, public health drop-in classes were closed indefinitely in Nova Scotia and public health visits turned virtual.^3^ This contrasted pre-pandemic support of visits with family and friends or attending new parent groups^4,5^ and in-person public health nurse visits.^6^ Not surprisingly, studies have found that COVID-19 has resulted in a significant increase in rates of perinatal depression and anxiety.^7,8^ High levels of social and family support are associated with greater maternal self-efficacy in first time mothers,^9^ yet this support may be missing during the pandemic due to physical distancing recommendations, potentially leading to low maternal self-efficacy.

Innovative solutions to bridge the postpartum care gap, such as mobile health (mHealth), are more important than ever. The benefits of mHealth include personalized information, the ability to connect with others, receiving reassurance, and monitoring changes.^10,11^ An example of this is text4baby, a perinatal text message intervention in the United States, covering topics between pregnancy and the first year, including nutrition, flu prevention, mental health, breastfeeding, and immunization.^12^ Text4baby has shown promise in targeting perinatal women living in low-income to encourage adaptation of health behaviours,^13,14^ however, such a perinatal program does not exist in Canada. Thus, our team developed *Essential Coaching for Every Mother*, a text message program designed to send daily messages during the immediate six-week postpartum period.^15^ *Essential Coaching for Every Mother* was designed prior to COVID-19 but had not yet been evaluated. With the outbreak of COVID-19 and the opportunity to fill the sudden gap in postpartum education and support, the program was modified and offered immediately. The primary objective of this study is to evaluate the preliminary impact of *Essential Coaching for Every Mother* on maternal self-efficacy, social support, postpartum anxiety, and depression. The secondary objective was to explore the acceptability of the *Essential Coaching for Every Mother* program (COVID-19 stream).

## Methods

### Study Population & Sample Size

At IWK Health, approximately 169 primiparous women give birth each month.^16^ As this was part of a feasibility study, no power calculations were conducted. Instead, the target was to recruit 15% of primiparous women who gave birth over a three-month period (n=75 from an estimate of 500 expected births). First time mothers were targeted to minimize the confounding factor of parity.^17,18^

To participate, women must (1) have given birth to their first baby at IWK Health and live in Nova Scotia; (2) have daily access to a mobile phone with text message capabilities; (3) be over 18 years of age; and (4) speak and read English. Women were excluded if their baby was over 21 days old at time of recruitment. No exclusions were made for mothers of infants who required care in a neonatal intensive care unit or if their infant had any major abnormalities.

### Design

This was a prospective pre-post study to pilot the *Essential Coaching for Every Mother* program with first time mothers in Nova Scotia, Canada between July 15th and September 19th, 2020.

### Procedures

The recruitment process was reported elsewhere.^19^ Briefly, women were recruited via social media, news, and study posters in the hospital and could indicate interest in participating between 37 weeks’ gestation and birth (considered antenatal recruitment) or could enrol after they gave birth up to three weeks postpartum (considered postpartum recruitment). *Essential Coaching for Every Mother* messages are sent based on delivery date, with messages designed to start the evening of the second day after birth. Participants could enrol up to 21 days postpartum and participants who enrolled late did not receive messages programmed to be sent earlier in the program.

Surveys were collected at two time points – enrolment (shortly after birth) and after the program was completed (six-weeks postpartum). Women were able to message “STOP” to withdraw from the study at any time at which point they stopped receiving messages and were no longer asked to complete surveys.

### Intervention

*Essential Coaching for Every Mother* is a six-week postpartum text message program that was developed in consultation with postpartum mothers and healthcare providers with the goal of improving women’s psychosocial outcomes in the immediate postpartum period.^15^ *Essential Coaching for Every Mother* (COVID-19 stream) contained 56 messages related to newborn care and maternal mental health, with nine messages modified to be consistent with postpartum care provided during COVID-19 and five COVID-19 specific messages added. Feedback on the modifications was obtained from three mothers and seven healthcare providers who participated in the original development.^15^ Table 1 lists the topics covered in both the original program as well as the COVID-19 stream. *Essential Coaching for Every Mother* sends twice daily messages during the first two weeks, and then one daily message for the following four weeks.^15^ In the COVID-19 stream, there were three days in the first two weeks in which a third message was sent. Women self-selected preference for breastfeeding or formula feeding messages which they were able to modify. While the messages were only one-way, they were personalized with the mothers’ name, the infant’s name, and the infant’s gender. Additional resources were often offered as part of the texts for participants if they desired more information on a topic. The TextIt platform was used to program and schedule the messages and the Twillo gateway service was used to send and receive messages to participants.^20,21^

**Table 1.** Topics covered in Essential Coaching for Every Mother – COVID-19

| Essential Coaching for Every Mother –<br>Original version (n=53) | Essential Coaching for Every Mother –<br>COVID-19 version (n=56) |
| --- | --- |
| Anxiety (3)<br>Depression (3)<br>Maternal self-care (5)<br>Postnatal follow-up (4)<br>Breastfeeding (5)/Formula (5)<br>Feeding (4)<br>Infant concerns (5)<br>Cord care (3)<br>Well-baby care (6)<br>Normal development (6)<br>Crying (5)<br>Safe sleep (4) | Anxiety (3)*<br>Depression (2)<br>Maternal self-care (5)*<br>Postnatal follow-up (4)***<br>Breastfeeding (5)**/Formula (5)<br>Feeding (4)<br>Infant concerns (4)<br>Cord care (3)<br>Well-baby care (6)*<br>Normal development (6)<br>Crying (5)<br>Safe sleep (4)<br>COVID-19 (5) |

\* Contains an edited message related to COVID-19

### Measures

Background maternal demographics and newborn characteristics were collected at enrolment. At six-weeks, a survey collected data on whether mothers had any postnatal healthcare contacts or concerns for themselves or their infant. At both time points, participants completed standardized assessments of self-efficacy (primary outcome) as well as social support, postpartum anxiety, postpartum depression, and COVID-19 anxiety and stress (secondary outcomes). Participants were also asked to rate how beneficial they found the *Essential Coaching for Every Mother* program on the above psychosocial outcomes on a 5-point scale and completed an evaluative component on their experience with the program.

Maternal self-efficacy was measured using the Karitane Parenting Confidence Scale (KPCS) which is a 15-item tool to assess perceived self-efficacy of mothers of newborns.^22^ A cut-off score of 39 or less (out of a possible 45) is considered to be a clinically low perceived parenting self-efficacy with scores below 31 considered to be in the moderate or severe range.^22^

Social support was measured through the Multidimensional Scale of Perceived Social Support (MSPSS).^23^ This scale is a measurement of how much support a parent feels they get from family, friends and significant others from which subscales from each can be calculated as well as a total score.^23^

Postpartum anxiety was measured using the Postpartum Specific Anxiety Scale (PSAS).^24^ The PSAS is a valid and reliable measure to assess anxiety during the first six-months postpartum.^24^ The State-Trait Anxiety Inventory (STAI)^19^ was used to measure general anxiety; STAI-State has 20 items for assessing state anxiety, with higher scores indicating greater anxiety.^25^

Postpartum depression was measured using the 10-item Edinburgh Postnatal Depression Scale (EPDS).^26^ This scale can be used to screen mothers at risk for developing postpartum depression, with a score above 14 (out of 30) indicating a likelihood of having or developing postpartum depression.^26^

Anxiety specific to COVID-19 was measured using the Coronavirus Anxiety Scale (CAS).^27^ This is a 5-item scale developed specifically to be a brief mental health screener to identify probable cases of dysfunctional anxiety associated with the COVID-19 crisis.^27^ The Impact of Stressful Event Scale – Revised (IES-R)^28^ was used to measure the impact of COVID-19 related stress. This 22-item self-report scale assesses subjective distress caused by traumatic event, which for our purposes was the COVID-19 pandemic. The IES-R yields a total score (ranging from 0 to 12) and subscale scores can also be calculated for the Intrusion, Avoidance, and Hyperarousal subscales (ranging from 0-4).^28^

### Analysis

Data were analysed using Statistical Package for Social Sciences version 26. Demographic characteristics are expressed in means and standard deviations and percentages, as applicable. A p-value of 0.05 was considered significant for all outcomes. For the first objective, paired t-tests were used to determine changes from baseline to follow-up. As this was an exploratory study, consideration of covariates occurred (i.e., number of messages received, perinatal timing of enrolment, maternal age). Participants who had incomplete paired data (e.g., missing a fully complete questionnaire at one of the time points) were excluded from the analysis on that outcome. For the second objective, thematic analysis was used to analyse qualitative responses, which was led by the first author.^29^

## Results

### Participants

A total of 96 participants enrolled in the study, with 200 potential participants initiating contact, 70% antenatally.^19^ Overall, 95 were not eligible, nine were not interested, four withdrew after enrolling, and four did not complete the baseline survey. Out of the 88 remaining participants, 76 (86.4%) fully completed both surveys, 8 (9.1%) had partial baseline and 8 (9.1%) had partial or incomplete follow-up survey data.

Women were on average 30.81 years of age (standard deviation [SD]=4.7). Most identified as heterosexual (91.3%), white (87.5%) and were married (67.5%) or common-law (26.3%). Over half (51.8%) had a household income over $100,000CAN. Almost all women had singleton births (98.8%). Women were on average 39.3 weeks pregnant when they gave birth (SD=1.5 weeks, range=33.5-42 weeks). For 87.5% of participants, this was a planned pregnancy, and 67.5% had a vaginal birth, 32.5% had caesarean sections. Thirty percent had a history of diagnosed anxiety or depression.

Of the 88 participants enrolled, 42 (47.7%) received the full 56 messages. Women who enrolled late received on average 45.36 messages (SD=9.9 messages, range=27-55). Seventy-eight participants (88.6%) opted in for breastfeeding messages, with two participants changing to formula. The average time between completion of the baseline survey and follow-up survey was 36.8 days or 5.25 weeks (SD=6.28 days, range=22-53 days).

### Psychosocial Outcomes

Changes in psychosocial outcomes are shown in Table 2. While there were some significant correlations, when examining the scatterplots, no clear relationship of the covariates could be determined, and thus no covariates were considered in the analysis. Overall, self-efficacy improved between baseline and follow-up and anxiety measured using the STAI-State showed a significant decline. No significant changes over time were found across the other outcome measures (social support, postpartum specific anxiety, postpartum depression, COVID-19 related stress or anxiety).

**Table 2.**
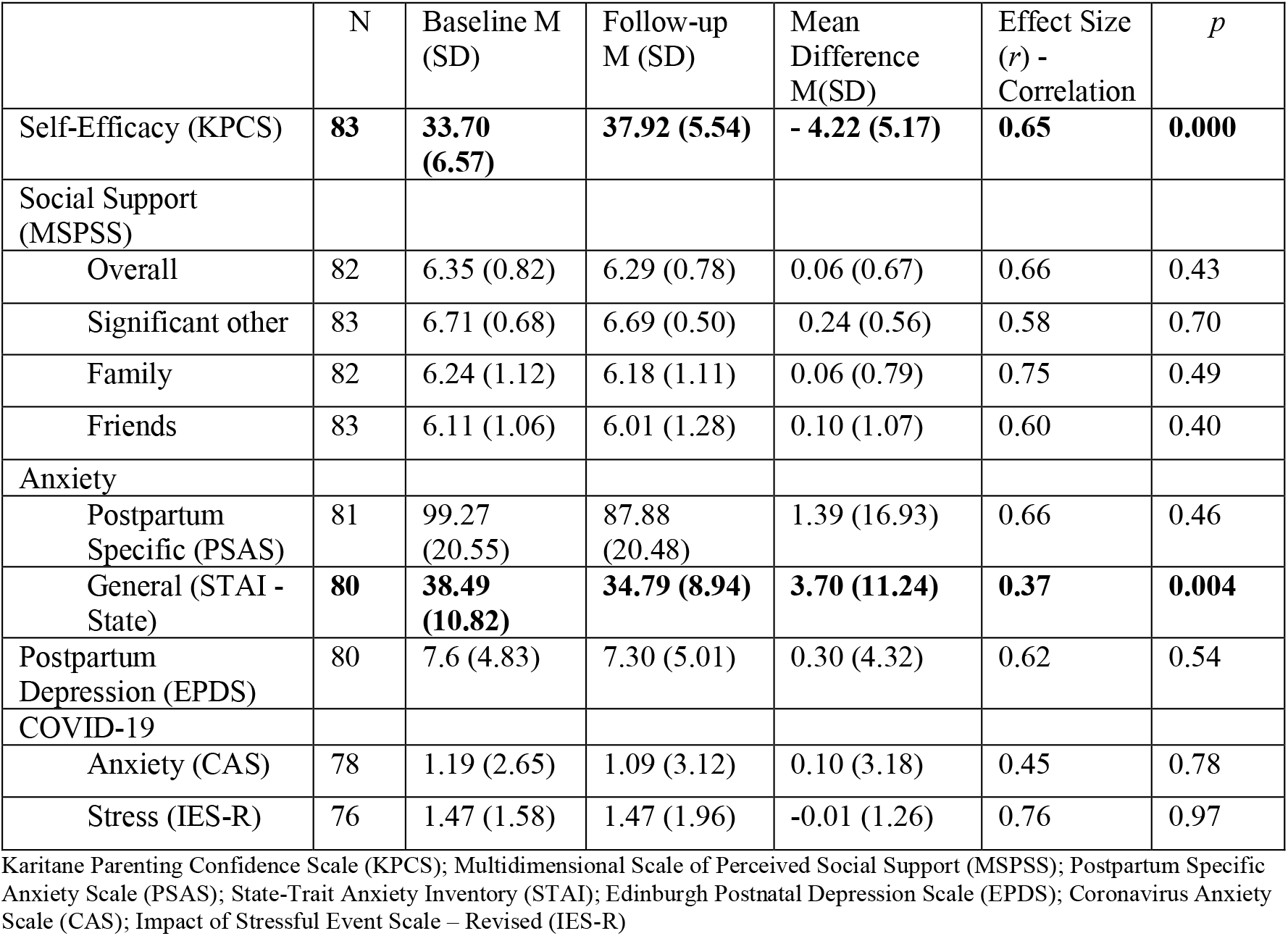
Changes in Psychosocial Outcomes from Baseline to Follow-Up

### Acceptability

Most participants (89.0%) felt that the number of messages were just right while the rest felt there were too few (7.3%) or too many (3.7%). Most (84.5%) felt the messages contained all the information they needed related to caring for a newborn and 98.8% indicated they would recommend the program to other new mothers. Table 3 shows how helpful participants found *Essential Coaching for Every Mother* in relation to the psychosocial outcomes measured on a scale from 1 (not at all) to 5 (completely).

**Table 3.** Participant self-reported rating of benefit of *Essential Coaching for Every Mother* on Psychosocial Outcomes

|  | N | Rating<br>M (SD) | Rating<br>Median (range) |
| --- | --- | --- | --- |
| Overall satisfaction | 82 | 4.45 (0.58) | 4.68 (2.4 - 5) |
| Self-Efficacy | 82 | 3.95 (1.10) | 4.00 (0.1--5.0) |
| Social Support (overall) | 82 | 3.63 (1.31) | 3.90 (0.00-5.0) |
| Anxiety (overall) | 82 | 3.30 (1.34) | 3.65 (0.00-5.0) |
| Postpartum Depression | 82 | 2.83 (1.61) | 2.85 (0.00-5.00) |
| COVID-19 Anxiety | 83 | 2.72 (1.57) | 2.65 (0.00-5.00) |
| COVID-19 Stress | 83 | 2.79 (1.53) | 2.65 (0.00-5.00) |

Areas where participants wanted more information were primarily related to breastfeeding and newborn sleep. Participants generally felt the timing of messages was relevant to their newborn’s development. One mother said: “The messages I received were so relevant to the stage we were in! Sometimes I felt as though someone was listening to my conversations. Very, very helpful.” A few women did comment that the messages arrived after they had already experienced that concern, but the majority considered the timing appropriate.

Women felt that the messages were helpful, particularly in light of the reduced face-to-face healthcare provider support available during COVID-19 pandemic restrictions. Participants also indicated that the messages would be helpful even beyond COVID-19 and they felt re-assured in the information they were getting: “It made me feel more confident about the information I was obtaining rather than what I would google.” However, some participants felt that due to the low COVID-19 prevalence in Nova Scotia at the time of the program implementation, they had low COVID-19 anxiety and stress already, thus the COVID-19 messages were not that helpful and increased their anxiety. One participant explained: “Mainly because I didn’t have any concerns about COVID-19, I found those messages to be the least helpful, but I think they were still important.”

Participants reported feeling as though they had access to a daily digital nurse: “It was nice just to be having constant communication. Our families both live outside the Atlantic bubble, so we haven’t had the initial support that we had always envisioned. Having the texts felt like we were being cared for and gave advice that we imagined our parents would give.” Women indicated that the messages made them feel less alone, particularly in relation to the loss of family support and peer support parenting groups. Another aspect of the program that participants appreciated was the normalization of what to expect with their newborn. One participant commented: “Messages of the witching hour normalized our witching hour and I felt less concerned that something was wrong and more confident that this was normal.” Another participant explained: “There were several occasions when we were experiencing something, and then the text would indicate this was a normal time to experience that thing. It made me feel like we were hitting milestones (or challenges) as we should. There also was a feeling of togetherness.”

Overall, what participants felt what they liked most was the consistency of the daily messages and the personalization with their child’s name. One mother said: “I liked the consistency of the texts and looked forward to receiving them”. Participants felt that the information validated what they already knew and/or what they were already doing. The information was consistent with messaging they were receiving from other healthcare sources. Participants appreciated that the information came directly to them, that they did not have to seek it out and that they could trust the information provided.

Areas for improvement included enhancing the ability to reply and/or engage with the messages or a support person with whom they could ask additional questions or to seek further classification. A few women commented that being able to tailor or opt in for additional messages would have been helpful: “… tailoring it to responses of mothers if they have certain issues [that] they would like more support with [such as] breastfeeding or colic.” Many women suggested the *Essential Coaching for Every Mother* continue beyond the first six-weeks postpartum: “Would love for it to go on longer. Even if the texts came less often but continued until 6 months. At 6 weeks I feel I’ve figured some out but … I still have lots of questions and know we have lots ahead of us to figure out with months 4-5…”. Women also suggested that the program be available for their partner, as one participant said: “I ended up sending him a screen shot of the messages most days”.

## Discussion

The primary objective was to evaluate the preliminary impact of *Essential Coaching for Every Mother* on maternal self-efficacy, social support, postpartum anxiety, and postpartum depression. This study found that maternal self-efficacy increased, with an improvement from the moderate clinical range (i.e., a score of 31 to 35 out of 45) to the mild clinical range (i.e., 36 to 39 out of 45), and postpartum anxiety measured using the STAI-State decreased between baseline and the 6-week follow-up. However, there were no significant changes on feelings of social support, postpartum specific anxiety, postpartum depression, or COVID-19 anxiety and stress. The second objective was to explore the acceptability of the *Essential Coaching for Every Mother* program provided during COVID-19. Overall, participants were satisfied with the program, with almost all participants willing to recommend the program to other mothers. While there was room for improvement, there were aspects of the program that women appreciated, including the daily messages with information they knew they could trust, the general appropriateness of the timing of the messages in relation to their newborn’s age, and the support and normalization of the postpartum experience the messages provided.

Although it is encouraging that self-efficacy increased and anxiety decreased between baseline and six-weeks postpartum, it is difficult to delineate the impact of *Essential Coaching for Every Mother* above and beyond changes that can occur as women become more comfortable in their parenting role. Interestingly, a decrease in anxiety measured using the STAI-State was found yet there was no significant difference when measured using the postpartum-specific anxiety measure. The STAI-State asks general questions around anxiety such as feeling calm or secure within the past seven days,^33^ whereas the PSAS is very specific to the postpartum period, asking questions around worry about baby’s weight or bonding.^24^ Thus, these measurement tools may be getting at different aspects of anxiety in the postpartum period. The challenge is that there is no consensus to date on how to best measure postpartum anxiety.^34^ A systematic review on anxiety measurement tools used in the perinatal period found the most common measurement tools were the General Health Questionnaire, the STAI, and the Hospital Anxiety and Depression Scales with significant heterogeneity of measurement tools across studies.^34^ On the other hand, Dennis and colleagues^35^ found that most studies used the STAI to assess postpartum anxiety in their systematic review on postpartum anxiety prevalence. One challenge is that measurement tools used to measure postpartum anxiety were not specifically designed for this purpose.^34^ Further work is needed to understand and identify the best approach to measuring postpartum anxiety.

Given the emerging evidence around increased postpartum depression and perinatal anxiety in women during the COVID-19 pandemic,^7,8^ it is important to consider the impact and opportunity of virtually delivered interventions. In terms of acceptability of the *Essential Coaching for Every Mother* program, almost all women felt the program was acceptable in terms of timing and content and was recommendable to other women. Women felt supported in receiving the personalized messages and even desired the program to continue beyond the six-weeks of the intervention period. Interestingly, while the messages were modified to provide some content related to COVID-19, due to the low prevalence of the virus in Nova Scotia while the program was implemented, this had limited impact. Thus, further iterations of the program should be modified to reflect primarily changes in care, rather than repetition of recommendations around physical distancing and safety. This finding may have been different if implemented where epidemiology of the virus was higher. Nevertheless, the *Essential Coaching for Every Mother* program was well-received and should progress to evaluation in a randomized controlled trial (RCT) to determine true effectiveness on psychosocial outcomes. While it will be important for the RCT to determine initial effectiveness of the program on mothers in the first six weeks, further iterations of *Essential Coaching for Every Mother* could consider the expansion to partners, and beyond the initial six weeks.

Due to the flexible nature of remote recruitment, there was significant variation in when women enrolled in the program - with under half of the participants receiving the full slate of messages. However, when considering the impact of the number of messages missed, no clear relationship or trend could be determined. It did not appear that the number of messages missed played a significant role in influencing the psychosocial outcomes. For implementation as an RCT, a shorter postpartum enrollment window may be beneficial to increase the number of messages received as designed as well as ensure there is sufficient time gap between baseline and follow-up surveys.

### Limitations

While it is important to explore the potential impact of the program on participants, it is difficult to tease apart the impact of the program compared to the normal psychosocial adjustment that occurs during the postpartum period. Thus, the findings from this feasibility study should be interpreted with that lens. Additionally, not all participants completed the survey in full at both time points, thus there were differences in the sample size across each outcome variable. The survey may have been too time consuming, especially for new mothers, suggesting it is important to reduce the number of assessment tools used in future iterations to enhance completion rates.

## Conclusion

In conclusion, preliminary findings suggest that *Essential Coaching for Every Mother* may play a role increasing maternal self-efficacy and decreasing generalized anxiety, although future work with a control group is needed to delineate the true effect of the program beyond changes that occur in the postpartum period generally. Overall, mothers were satisfied with the *Essential Coaching for Every Mother* program and would recommend this to other mothers, during the COVID-19 pandemic and beyond. *Essential Coaching for Every Mother* holds potential to improve maternal outcomes in the immediate postpartum period.

## Data Availability

Data not available.

## DECLARATIONS

### Ethics approval

All procedures performed in studies involving human participants were in accordance with the ethical standards of the institutional research committee (IWK Health REB#1024984) and with the 1964 Helsinki declaration and its later amendments or comparable ethical standards.

### Consent to participate

Informed consent was obtained from all individual participants included in the study. All participants consented to the following statement: “I have reviewed all of the information in this consent form related to the study called: Feasibility of implementing Essential Coaching for Every Mother during COVID-19 and I have been given the opportunity to discuss this study with a research team member. I do not have any further questions.”

### Consent for publication

Additional informed consent was obtained from all individual participants for whom identifying information is included in this article. All participants who are quoted consented to the following statement: “I agree that direct quotes from my survey may be used without identifying me.”

### Availability of data and material

Due to the ethical approval obtained, data cannot be made available.

### Authors’ contributions

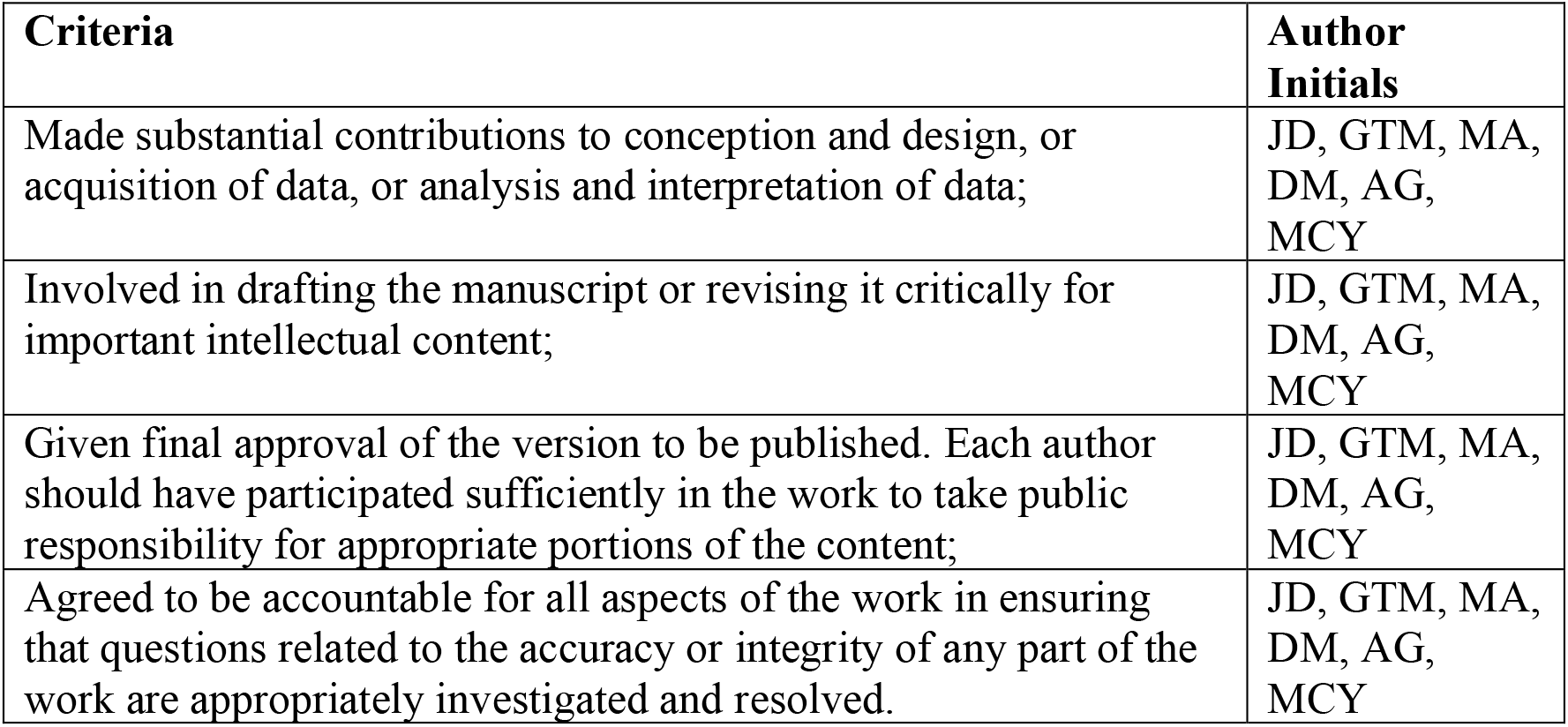

